# Genome-wide association study in 8,956 German individuals identifies influence of ABO histo-blood groups on gut microbiome

**DOI:** 10.1101/2020.07.09.20148627

**Authors:** Malte Christoph Rühlemann, Britt Marie Hermes, Corinna Bang, Shauni Doms, Lucas Moitinho-Silva, Louise Bruun Thingholm, Fabian Frost, Frauke Degenhardt, Michael Wittig, Jan Kässens, Frank Ulrich Weiss, Annette Peters, Klaus Neuhaus, Uwe Völker, Henry Völzke, Georg Homuth, Stefan Weiss, Matthias Laudes, Wolfgang Lieb, Dirk Haller, Markus M. Lerch, John F. Baines, Andre Franke

**Author notes:** **Correspondence should be addressed to:** Prof. Dr. Andre Franke, Institute of Clinical Molecular Biology (IKMB), Kiel University, Rosalind-Franklin-Str. 12, 24106 Kiel, Germany.

## Abstract

The intestinal microbiome is implicated as an important modulating factor in multiple inflammatory,^1,2^ neurologic,^3^ and neoplastic diseases.^4^ Recent genome-wide association studies yielded inconsistent, underpowered and rarely replicated results such that the role of human host genetics as a contributing factor to microbiome assembly and structure remains uncertain.^5–11^ Nevertheless, twin-studies clearly suggest host-genetics as driver of microbiome composition.^11^ In a genome-wide association analysis of 8,956 German individuals, we identified 32 genetic loci to be associated with single bacteria and overall microbiome composition. Further analyses confirm the identified associations of ABO histo-blood groups and FUT2 secretor status with *Bacteroides* and *Faecalibacterium*. Mendelian randomization analysis suggests causative and protective effects of gut microbes, with clade-specific effects on inflammatory bowel disease. This holistic investigative approach of the host, its genetics, and its associated microbial communities as a ‘metaorganism’ broadens our understanding of disease aetiology and emphasizes the potential for implementing microbiota in disease treatment and management.

## Main

We conducted the largest single country genome-wide association analysis of microbial traits followed by Mendelian Randomization (MR) analysis to elucidate the genetic link between humans and their associated microbiota. Our study comprised five independent cohorts from German biobanks located in Northern Germany (Kiel, Schleswig-Holstein; PopGen^12^, n=724; FoCus, n=957), North-Eastern Germany (Greifswald, Mecklenburg-Western Pomerania; SHIP, n=2,029; SHIP-TREND, n=3,382),^13,14^ and Southern Germany (Augsburg, Bavaria; KORA, n=1,864;^15,16^ see **Methods** for details).

Baseline comparisons show similarities in anthropometric measures, genomic variation and microbial community compositions between cohorts (**Figure 1, Supplementary Figure S1**, **Supplemental Material**). Taxonomic groups and sequence similarity clusters included in the univariate analysis, henceforth called microbial features, covered between 98.4% and 98.7% of the whole community at the phylum level and between 77.8% (PopGen) and 82.6% (SHIP-TREND) at the genus level across cohorts. These data indicate that the cohorts share a common core microbiota (cohort-level summaries of microbial features can be found in **Supplementary Table S1**).

**Figure 1:**
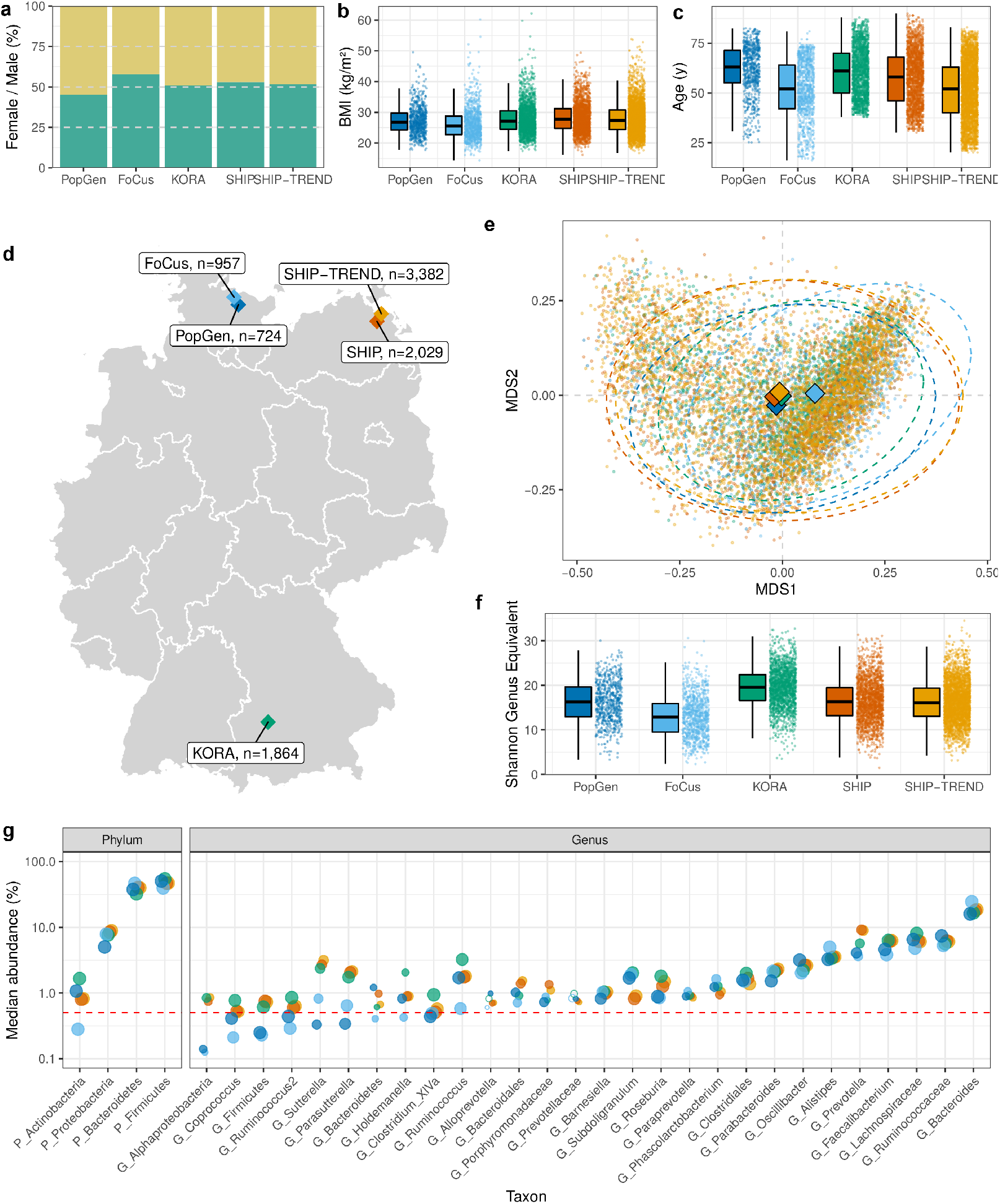
Summary of cohort properties. **(a)** Composition of study participants sex. **(b)** Distribution of participants BMI and **(c)** age. **(d)** Biobank/cohort locations in Germany. **(e)** Ordination of all samples based on genus-level Bray-Curtis dissimilarity. Diamonds represent cohort centroids, dashed ellipses represent 95% confidence level of multivariate t-distributions. (f) Distribution of alpha diversities as calculated by Shannon diversity genus-level equivalent and the number of observed genera. **(g)** Comparison of relative abundances of phylum- and genus-level taxonomic groups that met the inclusion criteria for the genome-wide association study in the five analysed German cohorts. Y axis represents the median abundance of the samples with non-zero abundance of the respective taxa, point size is relative to the prevalence of the respective taxon in the cohort. Taxa with cohort prevalence below the inclusion threshold of 20% are displayed as empty circles. The dashed red line represents the abundance threshold of .5% for inclusion in the analysis. Taxa are arranged from left to right by the lowest median abundance over all cohorts from high to low. Cohort-level summaries of microbial features can be found in **Supplementary Table S1**. In (b), (c) and (f), centre lines represent median values, box limits show 1^st^ and 3^rd^ quartile, whiskers extend to 1.5 interquartile ranges (IQR) ± 1^st^/3^rd^ quartile.

Univariate microbial features were defined based on taxonomic annotations from phylum to genus level. As taxonomic assignments below genus level don’t perform well,^17^ finer scale features were defined by sequence similarity clustering (97%- and 99%-similarity) and amplicon sequence variants (ASVs) to create a comprehensive dataset (see **Methods**). Host-features encoded by genetics can possibly influence presence-absence patterns of microorganisms, and also lead to shifts in the relative abundances of such, thus both assumptions were tested in the association analysis. In total, 198 and 233 univariate microbial features were analysed using logistic and linear regression, respectively (see **Methods** for details). Host-genetic variation might affect multiple community members, thus in addition to univariate analyses, whole community multivariate association analysis of genus-level Bray-Curtis dissimilarity and weighted UniFrac distance^18^ were performed (see **Methods**). Per-cohort results were combined in a meta-analysis framework (see **Methods**). To ensure robustness of results, genome-wide significant results (*p*_Meta_<5×10^-8^) were reported when supported by nominal significance (*p*<0.05) in at least two cohorts. Additionally, a study-wide significance threshold was defined as *p*_Meta_<1.866×10^-10^ and heterogeneity measures were calculated (see **Methods**).

Accordingly, we reveal a total of 44 genome-wide significant associations with microbial features and community composition involving 38 genomic loci (**Table 1**, **Figure 2a**), among which four associations stem from the multivariate analysis, 17 from the univariate abundance analysis, and 17 from the presence-absence patterns. The majority of genome association – including found in the presence/absence models – showed low heterogeneity (I^2^<40%), with only six abundance-associated variants showing moderate heterogeneity (I^2^<60%) and two surpassing this threshold, thus should be interpreted with caution. The top 10,000 genetic variants for univariate and multivariate analyses are summarized in **Supplementary Tables S2-4**. All results can be queried via the mGWAS results browser (http://ikmb.shinyapps.io/German_mGWAS_Browser). None of the signals surpassed the conservative threshold of study-wide significance. Univariate signals with overlapping genetic loci in all cases are found from the same taxonomic group at a different taxonomic and/or clustering level.

**Table 1:**
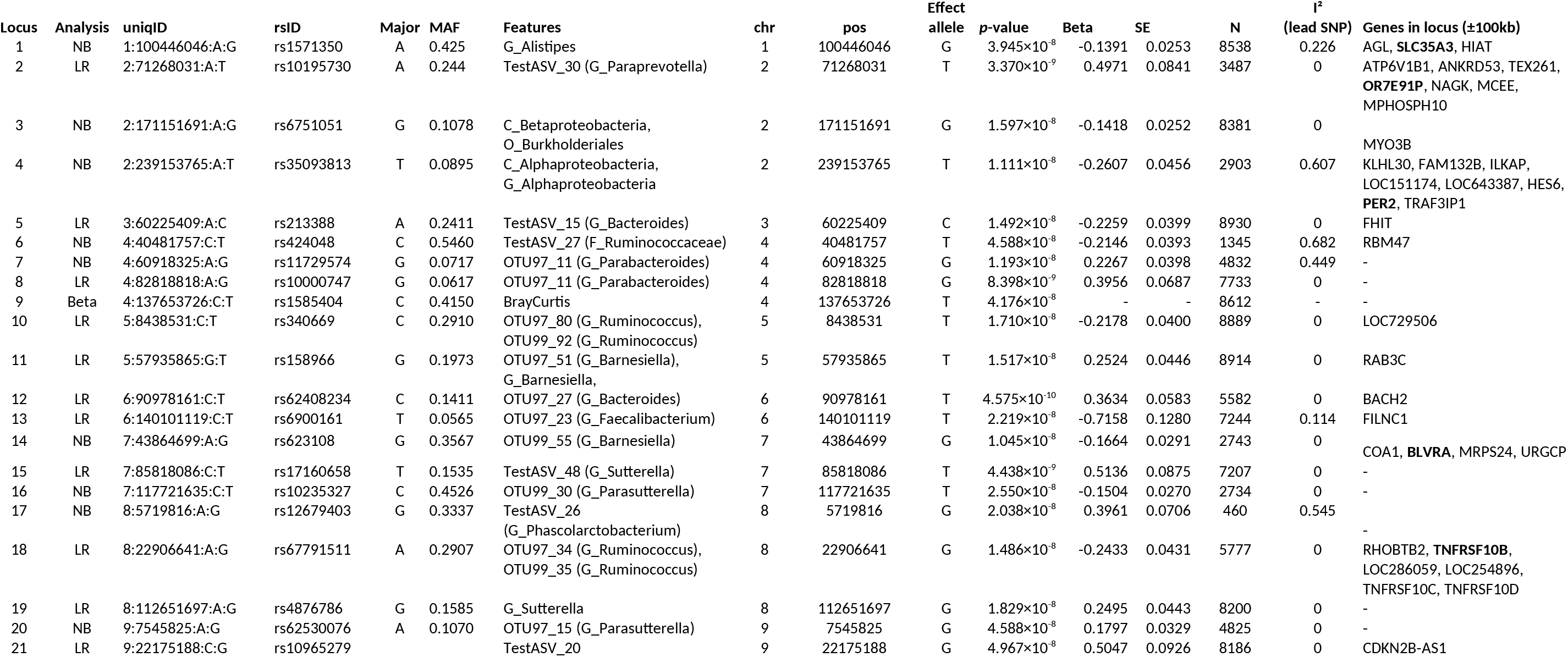
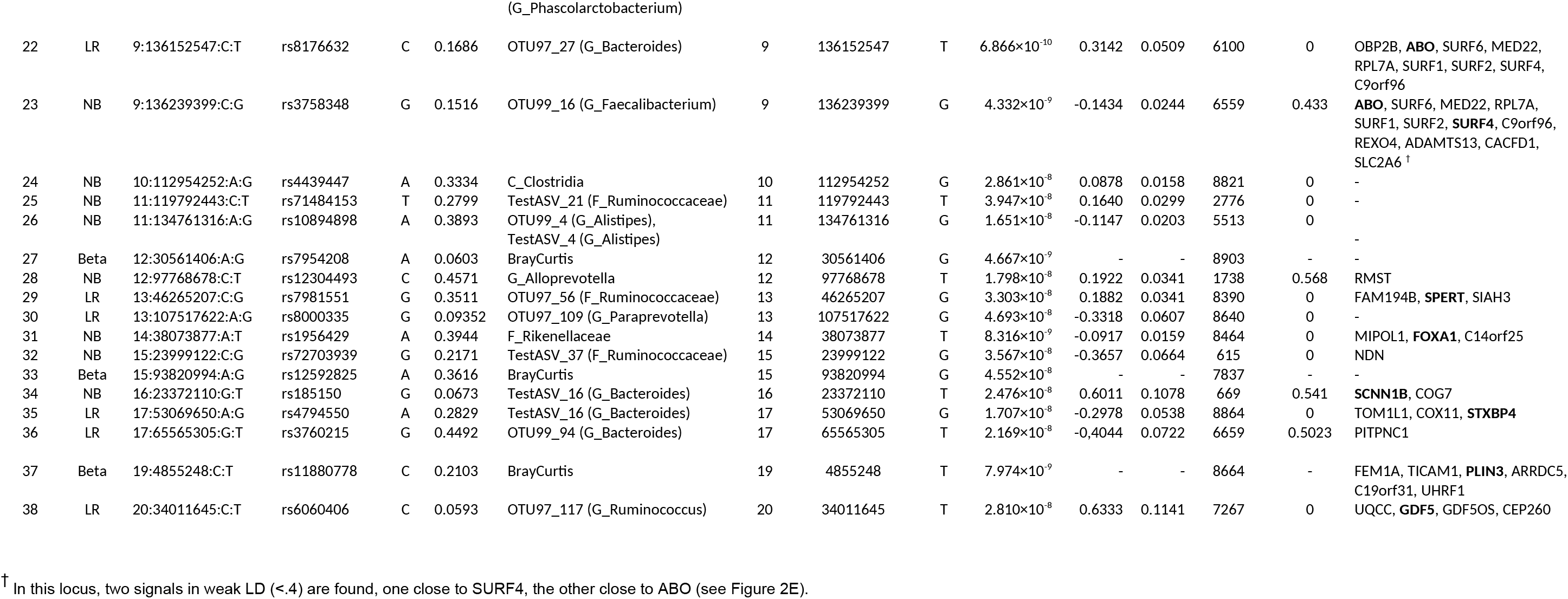
Results summary of the genome-wide association analysis for beta diversity (column “Analysis”: Beta), logistic regression of presence/absence patterns (LR) and analysis of abundances (NB) ordered and enumerated by genomic location of the loci. A single-variant association test was performed for each cohort and each microbial feature, adjusting the respective model for the first ten genetic principle components, age, sex and body-mass index (BMI). Results were meta-analysed weighted by inverse-variance for univariate and sample-size in multivariate non-parametric models. For univariate analysis, meta-analysis effect size (Beta) and standard error (SE) are given with respect to the effect allele, total sample numbers in the meta-analysis are given in column “N”. A genome-wide significance threshold of *p*_Meta_<5×10^-8^ and nominal significance (*p*<0.05) in at least two cohorts was considered to ensure robustness. I^2^ values for the lead SNPs are given as measure of heterogeneity. Genes up to 100kb up- and downstream of the lead SNP are listed, in case multiple genes are found in the locus, the closest gene to the lead SNP is marked in bold.

**Figure 2:**
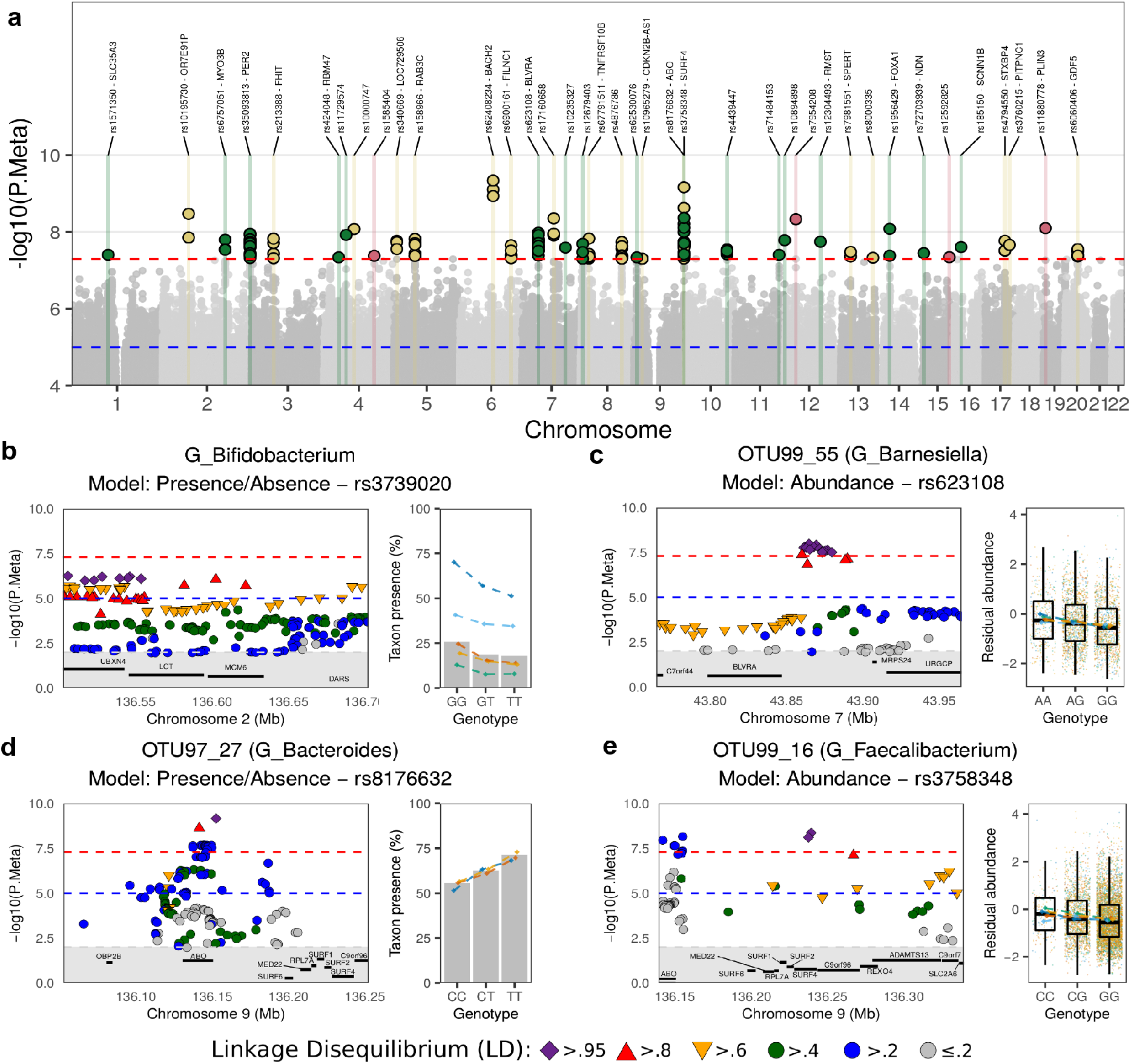
*Genome-wide association analysis results*. **(a)** Manhattan plot of *p*-values from the meta-analysis across all tested traits, the lowest *p*-value at each position is shown. Colour coding by analysis type. Green: abundance models; Yellow: presence-absence models (logistic regression); Red: beta diversity. Regional association plot of: **(b)** genus *Bifidobacterium* presence-absence test with variants in the *LCT* gene locus. **(c)** OTU97_55 *(Barnesiella)* abundance vs. variants at the biliverdin reductase A *(BLVRA)* gene locus. **(d)** OTU99_16 *(Faecalibacterium)* abundance vs. variants in the *ABO/SURF4* gene locus. **(e)** OTU97_27 *(Bacteroides)* presence-absence vs. *ABO* variants. The per-cohort feature abundance means and presences for each genotype are given by the diamonds in the respective colours. In panels (c) and (e) all residual abundance for the individual samples are displayed as dots in the respective colours of the cohort. Centre lines represent median values, box limits show 1^st^ and 3^rd^ quartile, whiskers extend to 1.5 interquartile ranges (IQR) ± 1^st^/3^rd^ quartile.

Although not meeting the initial inclusion criteria (see **Methods**), the genus *Bifidobacterium* was included in the analysis. Its connection with the lactase gene locus *(LCT)* on chromosome 2 is important, as it is the only signal replicating across numerous previous studies.^5,9,11^ The meta-analysis shows a clear association peak in the *LCT* locus with 53 variants displaying *p*-values lower than the suggestive *p*<10^-5^ threshold, the lowest for rs3820794 (chr2:136505546; *p*_Meta_= 5.62×10^-7^; **Figure 2b**). This is supported by nominally significant *p*-values in four of the five cohorts, with only the FoCus cohort showing a *p*-value above nominal significance (*p*_Focus_= 0.069), underlining the previously found connection between the *LCT* locus and *Bifidobacterium* and the validity of the herein-used model of choice, although LD structure does not pinpoint the *LCT* gene itself as the location of the primary association. Also, connections to age and consumption of dairy products remain unresolved and need to be investigated through more targeted approaches.^11,19^

Our obtained genome-wide association results point to immune-mediated interactions of host and microbiota, e.g. the association detected for OTU99_55 *(Barnesiella;* OTU: operational taxonomic unit) and variants in the biliverdin reductase A *(BLVRA;* rs623108; *p*_Meta_=1.05×10^-8^; **Figure 2c**) locus. Biliverdin reductase A was previously shown to inhibit Toll-like receptor 4 (TLR4) gene expression.^20^ TLR4 is a pattern recognition receptor that initiates an immune response to bacterial lipopolysaccharides (LPS) present in many Gram-negative bacteria.^21^ *Barnesiella*, which itself is Gram-negative, is negatively associated with LPS-induced interferon-gamma production, suggesting a contribution of this commensal to homeostasis by immune- or TLR4-signal-modulation.

We identified two independent univariate associations with a locus surrounding the histo-blood group ABO system transferase (ABO) gene. One *ABO* gene signal for differential abundance includes OTU99_16 belonging to *Faecalibacterium* (rs3758348; chr9:136155000; *p*_Meta_=6.16×10^-9^; **Figure 2d**), which is accompanied by a second signal ~100kb downstream in the surfeit locus protein 4 *(SURF4)* gene (chr9:136239399; *p*_Meta_=4.33×10^-9^). The second *ABO* association is between rs8176632 allele T and the increased prevalence of a *Bacteroides* OTU (OTU97_27; rs8176632; chr9:136152547; *p*_Meta_=6.87×10^-10^; Figure 2E). Interestingly, this same *Bacteroides* OTU is also significantly associated with variants at the *BACH2* (BTB domain and CNC homolog 2) gene locus (chr6:90978161; *p*_Meta_=4.58×10^-10^). Moreover, a suggestive association between this *Bacteroides* OTU is present for the *FUT2* (Galactoside 2-alpha-L-fucosyltransferase 2) locus, whereby the strongest signal is from the missense variant rs602662 (chr19:49206985; *p*_Meta_=4.46×10^-7^), which is in strong linkage disequilibrium (LD) with variant rs601338 (R^2^=0.8898) encoding the FUT2 secretor phenotype. This variant determines whether the fucosyl-precursor for the ABO blood-group system is synthesized on mucosal surfaces in the body and secretions. Individuals homozygous for this missense variant do not have the ABO-encoded antigen on mucosal cells, independent of the ABO allele (i.e. display the non-secretor phenotype; **Figure 3b-d**). Variants at *FUT2* and *BACH2*, correlated with *Bacteroides* OTU97_27 in this study, were previously shown to be associated with inflammatory bowel disease (IBD).^22-25^

**Figure 3:**
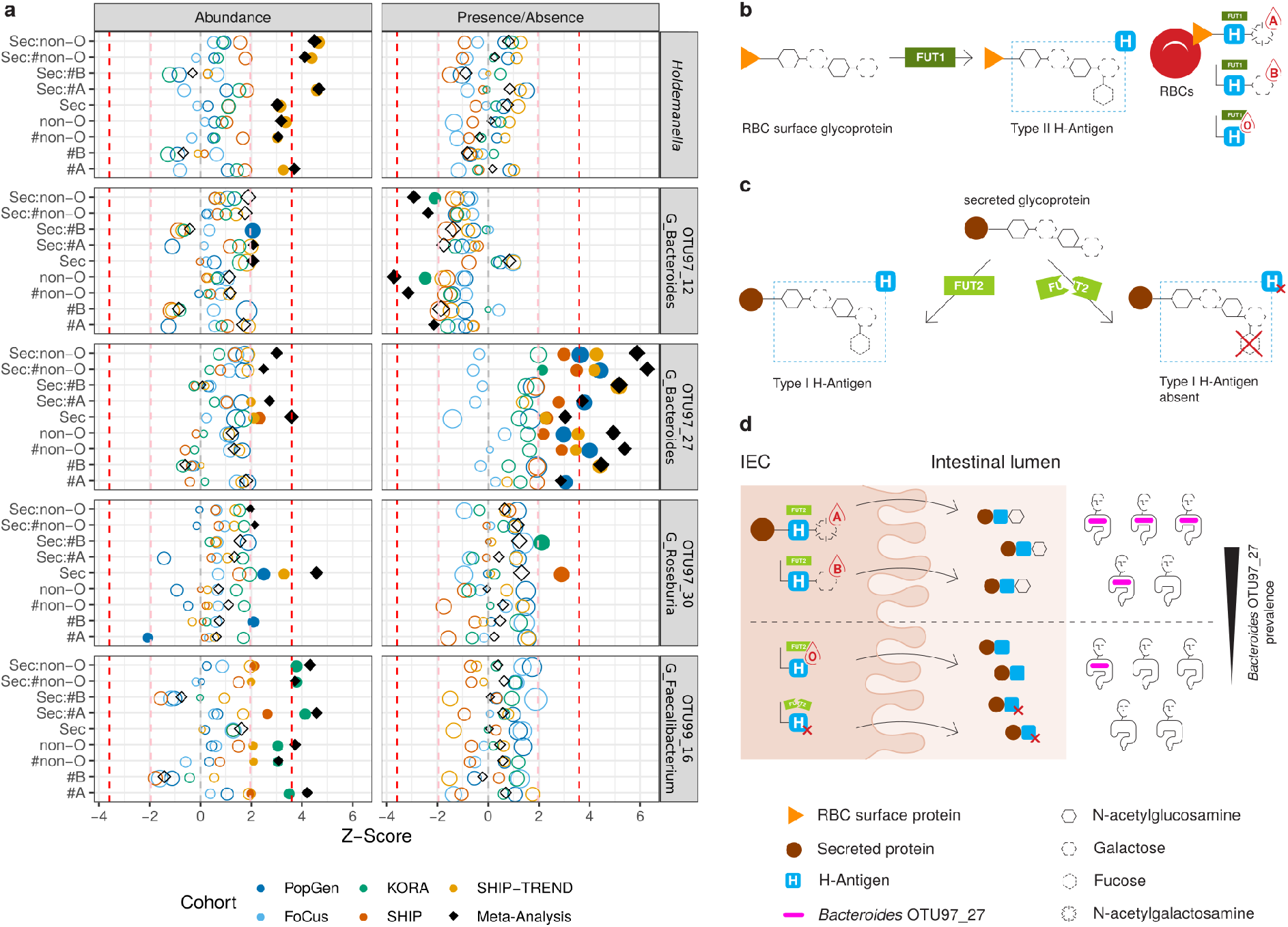
**(a)** Results of the analysis of nine models connecting feature prevalence and abundance to ABO blood group alleles and FUT2 secretor status. Shown are all univariate microbial features with at least one meta-analysis *q*-value < 0.05. *Holdemanella* is shown also representing *Holdemanella* OTU97_33, and *Bacteroides* OTU97_12 is shown representing also OTU99_12 and TestASV_13 of the same *Bacteroides* subclade with respective identical results. The Y-axes represent the nine models applied, investigating the linear effects of the number of A (#A) and B (#B) histo-blood group alleles and their sum (#non-O), as well as the effects of binary traits O vs. non-O histo-blood group (non-O) and FUT2 secretor status (Sec). The statistical interaction of Sec with all former traits is also included, indicated by the colon (:) symbol; The X-axis shows the Z-scores of the respective models. Symbols are coloured according to cohort, black diamonds represent the result of the meta-analysis of all five cohorts. Symbol size represents absolute effect size. *p*-values < 0.05 are displayed as solid shapes. Dashed vertical lines represent Z-values corresponding to nominal significance (light red line: *p*<0.05; Z=±1.96; two-sided) and adjusted significance (dark red line: *q*<0.05; Z=±3.59; two-sided). The complete results can be found in **Supplementary Table S5**. **(b)** The type II H-antigen on red blood cells (RBCs) is completed by addition of a fucose sugar by the enzyme Fucosyltransferase 1 (FUT1). Subsequently the A- and B-antigens are synthesized by addition of N-acetylgalactosamine or galactose, respectively. In individuals with a O histo-blood group, no additional sugars can be added to the H-antigen. **(c)** On secreted proteins and mucosal cells, the fucosylated type I H-antigen is synthesized by the enzyme Fucosyltransferase 2 (FUT2). In individuals homozygous for the rs601338-A missense variant in *FUT2* – also known as non-secretors – there is no addition of a fucosyl-group, resulting in no H-antigen. **(d)** Consequently, no additional sugars are added to the precursor of the H-antigen in non-secretors, irrespective of the individuals’ histo-blood group genotype at ABO. *Bacteroides* OTU97_27 exhibits higher prevalence in individuals with non-O ABO histo-blood groups and functioning FUT2 as compared to individuals with O histo-blood group or FUT2 non-secretors.

For a focused evaluation of blood-group dependent associations with microbial features, we investigated ABO histo-blood group and FUT2 secretor status (see **Methods**). The prevalence or abundance of eight taxonomic groups show at least one FDR-corrected significant association (q<0.05) with either ABO histo-blood group alleles, secretor status, or their interaction (**Figure 3a**; **Supplementary Table S5**). These results demonstrate a positive correlation between non-O blood group and positive secretor status and the prevalence of the aforementioned *Bacteroides* OTU97_27 in four of the five cohorts (*p*_Meta_=3.65×10^-10^). Intriguingly, a different *Bacteroides* branch, represented by OTU97_12, OTU99_12, and TestASV_13, exhibited significant associations with ABO histo-blood group status as well, however in this case characterized by an inverse relationship between prevalence and non-O blood group alleles (*p*_Meta_=2.1×10^-4^). Together, these findings suggest histo-blood group dependent effects on *Bacteroides* subclades.

In addition, the model points to an association between *Faecalibacterium* OTU99_16 and the ABO histo-blood group A allele in interaction with secretor status (*p*_Meta_=4.7×10^-6^). A significant association between *Holdemanella* and ABO is also identified, although the signal is exclusively driven by the SHIP-TREND cohort with only weak support from the remaining cohorts. Further, FUT2 secretor status is associated with differential abundance of *Roseburia* OTU97_30, independent of ABO blood type (*p*_Meta_=4.79×10^-6^). In conclusion, the analyses reveal a specific impact of the human ABO blood groups and secretor status on members of the intestinal community.

Mendelian randomization (MR) has recently become a popular tool to infer causal relationships of complex traits in observational data,^26^ and recent publications suggest that MR can be used for exploratory inference of causal effects the microbiome may have on complex host traits.^27^ MR analysis was performed for all univariate microbial features as "exposures” and 41 selected binary traits from the MR-Base database^28^ as outcomes (see **Methods**). This allows us to assess potential causal effect of microbial features on disease. A total of 19 comparisons reach the per-trait suggestive threshold of *p*<1.22×10^-3^, with five traits falling below the global FDR-correction threshold q<0.05 (**Table 2**; **Supplementary Table S6**). Nine out of 19 suggestive microbial effects on host traits point to IBD and its sub-entity Crohn’s disease. For example, the presence of the same *Bacteroides* OTU associated with ABO histo-blood group status (OTU97_27) and a *Prevotella* ASV (TestASV_18) appear to significantly protect against CD development (β=-0.515 and β=-0.257, respectively). Previous work has revealed *Bacteroides-* and *Prevotella* as main determinants of gut enterotypes.^29-31^ Recent studies applying quantitative microbiome profiling suggested protective effects of *Prevotella-dominated* communities on CD, as well as contradicting connections of IBD to *Bacteroides-subclades*, potentially modulated by microbial load,^32^ supported by additional studies pointing at lower abundances of *Bacteroides* being associated with IBD development.^33^ These results once again emphasize the large variability of *Bacteroides* taxa in connection to genetics and disease.

**Table 2:** Results from Mendelian Randomization (MR) analysis. Shown are only results with *p*<1.220×10^−3^ (significance threshold as determined in **Methods**) and the respective FDR-adjusted *q*-values. All SNPs with F-statistics > 10 and *p*<10^-5^ in the respective genome-wide association meta-analysis of presence/absence (LR) and abundance (NB) patterns (exposures) were used as instrument variables and tested for their effects on 41 binary traits (see **Methods** and Supplementary Material). Mean and minimum F-statistics of included instruments are reported. Tests used for MR (Method) were Wald ratio (WR) in case of a single instrument variable, and inverse-variance weighted (IVW) analysis in case of two and more instrument variables (#SNPs). Effect sizes (Beta) and standard errors (SE) of the primary analyses are reported in the table. A complete table of all results from all MR analyses including results from the sensitive analysis can be found in **Supplementary Table S6**.

| Outcome | Exposure | Analysis | Method | #SNPs | F (Mean) | F (Min) | Beta | SE | p-value | q-value |
| --- | --- | --- | --- | --- | --- | --- | --- | --- | --- | --- |
| **Anthropometric** |  |  |  |  |  |  |  |  |  |  |
| Extreme body mass index \|\| id:85 | P_Firmicutes | NB | IVW | 2 | 20.17 | 18.81 | -0.8804 | 0.2634 | $8.31 \times 10^{-4}$ | 0.4044 |
| Obesity class 2 \|\| id:91 | G_Parabacteroides | NB | IVW | 3 | 18.32 | 18.09 | -0.5683 | 0.1659 | $6.14 \times 10^{-4}$ | 0.3350 |
| **Autoimmune / inflammatory** |  |  |  |  |  |  |  |  |  |  |
| Asthma \|\| id:44 | OTU99_84 (G_Prevotella) | LR | WR | 1 | 11.77 | 11.77 | -0.7256 | 0.2109 | $5.82 \times 10^{-4}$ | 0.3927 |
| Celiac disease \|\| id:1059 | OTU99_62 (F_Ruminococcaceae) | LR | WR | 1 | 21.22 | 21.22 | -0.6131 | 0.1763 | $5.07 \times 10^{-4}$ | 0.3764 |
| Crohn's disease \|\| id:10 | C_Clostridia | NB | WR | 1 | 19.28 | 19.28 | -1.7060 | 0.2465 | $4.46 \times 10^{-12}$ | $4.28 \times 10^{-8}$ |
| Crohn's disease \|\| id:10 | OTU97_27 (G_Bacteroides) | LR | WR | 1 | 17.45 | 17.45 | -0.5151 | 0.0867 | $2.77 \times 10^{-9}$ | $1.33 \times 10^{-5}$ |
| Crohn's disease \|\| id:10 | TestASV_23 (G_Barnesiella) | LR | WR | 1 | 15.17 | 15.17 | 0.2711 | 0.0599 | $6.00 \times 10^{-6}$ | 0.0115 |
| Crohn's disease \|\| id:10 | TestASV_18 (G_Prevotella) | LR | WR | 1 | 13.42 | 13.42 | -0.2575 | 0.0579 | $8.76 \times 10^{-6}$ | 0.0140 |
| Crohn's disease \|\| id:11 | F_Porphyromonadaceae | NB | IVW | 2 | 21.01 | 20.43 | 3.2134 | 0.7962 | $5.44 \times 10^{-5}$ | 0.0745 |
| Crohn's disease \|\| id:11 | TestASV_12 (G_Bacteroides) | LR | WR | 1 | 20.44 | 20.44 | -1.0344 | 0.3160 | $1.06 \times 10^{-3}$ | 0.5672 |
| Inflammatory bowel disease \|\| id:293 | F_Porphyromonadaceae | NB | IVW | 2 | 21.01 | 20.43 | 2.5143 | 0.5433 | $3.70 \times 10^{-6}$ | $8.88 \times 10^{-3}$ |
| Inflammatory bowel disease \|\| id:293 | TestASV_12 (G_Bacteroides) | LR | WR | 1 | 20.44 | 20.44 | -0.9989 | 0.2654 | $1.68 \times 10^{-4}$ | 0.1786 |
| Inflammatory bowel disease \|\| id:293 | OTU99_85 (G_Alistipes) | LR | WR | 1 | 17.18 | 17.18 | 0.4096 | 0.0776 | $1.29 \times 10^{-7}$ | $4.13 \times 10^{-4}$ |
| **Cancer** |  |  |  |  |  |  |  |  |  |  |
| Ovarian cancer \|\| id:1120 | OTU97_27 (G_Bacteroides) | LR | IVW | 5 | 14.54 | 13.44 | -0.1140 | 0.0341 | $8.39 \times 10^{-4}$ | 0.4740 |
| Gallbladder cancer \|\| id:1057 | OTU97_4 (G_Alistipes) | NB | IVW | 4 | 14.71 | 13.52 | 5.8987 | 1.5071 | $9.08 \times 10^{-5}$ | 0.1090 |
| **Cardiovascular** |  |  |  |  |  |  |  |  |  |  |
| Coronary heart disease \|\| id:6 | TestASV_23 (G_Barnesiella) | LR | IVW | 4 | 17.78 | 15.17 | 0.1497 | 0.0431 | $5.10 \times 10^{-4}$ | 0.3764 |
| **Psychiatric / neurological** |  |  |  |  |  |  |  |  |  |  |
| Autism \|\| id:802 | TestASV_11 (F_Lachnospiraceae) | NB | IVW | 7 | 11.44 | 10.60 | 0.4156 | 0.1151 | $3.07 \times 10^{-4}$ | 0.2945 |
| Major depressive disorder \|\| id:804 | OTU97_51 (G_Barnesiella) | NB | WR | 1 | 16.49 | 16.49 | 0.8655 | 0.2447 | $4.05 \times 10^{-4}$ | 0.3538 |
| Schizophrenia \|\| id:22 | F_Lachnospiraceae | NB | IVW | 8 | 21.45 | 19.96 | 0.1687 | 0.0521 | $1.20 \times 10^{-3}$ | 0.6069 |

Further results from MR confirm host-microbiome interactions previously described in observational studies. *Parabacteroides* show a protective effect on the "Obesity class 2” trait (β=-0.568), supporting previous experimental observations of *Parabacteroides* species alleviating obesity effects in mice.^34^ Interestingly, none of the microbial traits with causal effects reach genome-wide significance at any locus in the univariate analysis. In addition to MR, replication of previously associated loci and gene-set enrichment and tissue specificity analysis was performed using the FUMA web service^35^ (see **Supplemental Material**). The obtained results indicate metabolic interactions between the host and associated microbes and an enrichment of genes derived from metabolic and inflammatory traits.

Our results highlight the power of combining multiple independent cohorts for genomic association analyses of microbial features, as they allow for robust and replicable results. Although a direct influence of ABO histo-blood group and secretor status on the microbiome is debated,^36,37^ our results support this interaction, potentially acting as a modulator in diseases for which variants in histo-blood groups and the microbiome were independently reported as risk factors,^22,38-40^ The suggestive causative role of *Bacteroides* in patients genetically susceptible to IBD development is notable, as multiple independent, and sometimes contrasting, results were previously reported from host-microbe association and MR analyses. The multifaceted role of *Bacteroides* in the human gut microbiome is likely insufficiently captured by 16S rRNA gene amplicon-based surveys and may therefore require future in-depth strain-level analysis. Nevertheless, our results suggest an important role of the human ABO histo-blood group antigens as candidates for direct modulation of the human metaorganism in health and disease.

## Online Methods

### Cohort description, genotyping and imputation

**PopGen:** The PopGen cohort is a population-based cohort from the area around Kiel, Schleswig-Holstein, Germany.^12^ From this cohort, 1,108 individuals were genotyped using the Affymetrix Genome-Wide Human SNP Array 6.0 covering 906,600 genetic variants. After the initial QC, which included filtering out variants with a minor allele frequency (MAF) < 1%, per-SNP callrate < 95% and deviation from Hardy-Weinberg equilibirum (HWE) with *p*<10^-5^, the genotyping data were prepared for imputation following the miQTL cookbook instructions (https://github.com/alexa-kur/miQTL_cookbook#chapter-2-genotype-imputation). Briefly, this Plink-based processing script includes steps to prepare variants to be in consistency with the HRC v1.1 reference panel regarding the order of reference and alternative alleles, variant naming and strand orientation. Finally, all data is converted to VCF files for imputation. Imputation of the autosomal chromosomes was performed using the Michigan Imputation Server using the Haplotype Reference Consortium (HRC) release v1.1 from 2016 as reference panel. Eagle v2.3 was chosen as phasing algorithm and EUR individuals was selected as population for quality control purposes. The process was started in “Quality Control & Imputation” mode. After downloading the final data, it was converted to binary plink files. and variants with minor allele frequency < 1% were removed. Faecal samples were available for 724 of these individuals. Faecal samples were collected by the participants themselves at their respective home in standard faecal collection tubes and mailed to the study centre where they were stored at -80°C until processing. DNA from faecal samples (approx. 200 mg) was extracted using the QIAamp DNA stool mini kit automated on the QIAcube.

**Food Chain Plus (FoCus):** The FoCus cohort was incepted as part of the competence network Food Chain Plus (http://www.focus.uni-kiel.de/component/content/article/88.html). This cohort consists of two parts. One part is a population-registry based cross-sectional cohort including individuals from the area around Kiel, Schleswig-Holstein, Germany. The second part is an outpatient clinic-based cohort including obese individuals (BMI > 30) with and without accompanying disease status. For our study, only the registry-based part of the cohort was included. Cohort participants were genotyped using the Infinium OmniExpressExome array. Data processing, imputation and sampling of faecal material was performed in the same way as in the PopGen cohort. Finally, out of 1,583 participants, 957 belonged to the population-based part of the cohort and supplied faecal samples. DNA from faecal samples (approx. 200 mg) was extracted using the QIAamp DNA stool mini kit automated on the QIAcube.

**KORA FF4:** KORA (Kooperative Gesundheitsforschung in der Region Augsburg) is a population-based adult cohort study in the Region of Augsburg, Southern Germany, that was initiated in 1984 (https://www.helmholtz-muenchen.de/epi/research/cohorts/kora-cohort/objectives/index.html). For the second follow-up study (FF4) of baseline study S4 2,279 participants were recruited and the study was conducted in 2013/2014 mainly focusing on diabetes, cardiovascular disease, lung disease and links to environmental factors such as the microbiome. Stool-derived DNA samples of 2,136 participants were obtained via the KORA Biobank. The DNA had been extracted using a guanidinethiocyanat / *N*-lauroylsarcosine-based buffer^40^ and subsequent clean-up with NucleoSpin gDNA Clean-up (Macherey-Nagel) for further analysis. Genotyping was performed using the Affymetrix Axiom array, initial QC of raw data included MAF filtering < 1%, per-SNP callrate < 98% and deviation from Hardy-Weinberg equilibirum (HWE) with *p*<10^-4^-. In total, 1,864 samples with genotyping and 16S rRNA gene survey data were included in the association analysis.

**SHIP and SHIP-TREND:** The Study of Health in Pomerania (SHIP) is a longitudinal population-based cohort study located in the area of West Pomerania (Northeast Germany). It consists of the two independent cohorts SHIP (n = 4,308; baseline examinations 1997 – 2001) and SHIP-TREND (n = 4,420; baseline examinations 2008 – 2012 with regular follow-up examinations every five years.^13^ Stool samples have been collected since the second follow-up investigation of the SHIP (SHIP-2, 2008 – 2012) and the baseline examination of the SHIP-TREND cohort. All faecal samples were collected by the study participants in their home environment, stored in a plastic tube containing stabilizing EDTA buffer and shipped to the laboratory where DNA isolation (PSP Spin Stool DNA Kit, Stratec Biomedical AG, Birkenfeld, Germany) was performed as described before.^41^ For a total of 2,029 and 3,382 samples 16S rRNA gene survey and genotype on Affymetrix Genome-Wide Human SNP Array 6.0 and Illumina Infinium Global Screening Array, respectively, data were available and included in the association analysis. Initial QC of raw genotyping data included filter for per-SNP callrate < 95% and deviation from Hardy-Weinberg equilibirum (HWE) with *p*<10^-5^.

Written, informed consent was obtained from all study participants in all cohorts, and all protocols were approved by the institutional ethical review committee in adherence with the Declaration of Helsinki Principles.

### Inference of ABO blood group and secretor status

ABO blood groups were inferred using the phased and imputed genetic data and four variants as proposed by Paré *et al*.,^42^ which rs507666, rs687289, rs8176746, rs8176704 encode for the allele A1, O, B, and A2, respectively. All variants were, depending on the genotyping array used in the respective cohort, either genotyped by the array or showed very high imputation quality scores between 98.7% and 99.8%. Additionally, observed allele frequencies were manually compared to frequencies in public databases to assure highest quality blood group assignments. Secretor status was assessed by variant rs601338 on chromosome 19. Individuals homozygous for the A allele were classified as “non-secretor”. This variant was genotyped in all cohorts, except for the PopGen cohort. Here, the estimated imputation accuracy was 94.6%.

### Microbial data generation and processing

Library preparation and sequencing was performed using a standardized protocol at a single wet lab in Kiel, Germany. DNA amplification by polymerase chain reaction (PCR) of the bacterial 16S rRNA gene was performed using the 27F/338R primer combination targeting the V1-V2 region of the gene employing a dual-index strategy to achieve multiplex sequencing of up to 384 samples per sequencing run. After PCR, product DNA was normalized using the SequalPrep Normalization Kit. Sequencing of the libraries was performed on an Illumina MiSeq using v3 chemistry and generating 2×300bp reads. Demultiplexing was performed allowing no mismatches in the index sequences. Data processing was performed in the R software environment (version 3.5.1)^43^, using the DADA2 (v.1.10)^44^ workflow for big datasets (https://benjjneb.github.io/dada2/bigdata.html) resulting in abundance tables of amplicon sequence variants (ASVs). All sequencing runs underwent quality control and error profiling separately. Briefly, forward and reverse reads were trimmed to a length of 230 and 180 bp, respectively, or at the first position with a quality score less or equal to 5. Low quality read-pairs were discarded when the estimated error in one of the reads exceeded 2 or of ambiguous bases (“N”s) were present in the base sequence. Read pairs that could not be merged due to insufficient overlap or mismatches in their nucleotide sequences were discarded. The complete workflow adjusted for the 16S rDNA V1-V2 amplicon can be found on GitHub: https://github.com/mruehlemann/german_mgwas_code/tree/master/1_preprocess. Finally, all data from the separate sequencing runs were collected in a single abundance table per dataset, followed by chimera filtering. ASVs underwent taxonomic annotation using the Bayesian classifier provided in DADA2 and using the Ribosomal Database Project (RDP) version 16 release.^45^ ASV abundance tables and taxonomic annotation were passed on to the phyloseq package^46^ for random subsampling to 10,000 sequences per sample *(rarefy_even_depth())* and construction of phylum- to genus-level abundance tables *(tax_glom())*. Samples with less than 10,000 clean reads were not included in the analysis. Sequences that were not assignable to genus level were binned into the finest-possible taxonomic classification. As amplicon-based sequencing of the 16S rDNA has clade-dependent taxonomic resolution differences,^47^ abundance profiles of ASVs and operation taxonomic units (OTU) based on two widely used similarity cut-offs (97% similarity for a proxy of species level, 99% similarity for strain level) were included in the analysis. This enables for an unbiased assessment of genetic effects at a sub-genus taxonomic scale. Although similarity cut-offs as proxy for taxonomic resolution are element of ongoing discussion^48^, clustering still allows to bundle similar sequences, and by that evolutionary closely related organisms, into units of likely also functional similarity. For this, ASV datasets were exported including their respective abundance information and combined for a dataset-spanning OTU picking at 99% and 97% identity level using the VSEARCH software.^49^ ASVs and OTUs were assigned cross-dataset consistent IDs for more convenient data handling, 97%- and 99%- identity based features being named OTU97 and OTU99 throughout the article, respectively. ASVs included in the analysis were relabelled to “TestASV”. OTUs on 97% identity level were aligned against the SILVA reference alignment (v132) using the SINA aligner, consistent gaps in the alignment were truncated.^50^ The resulting alignment was used to construct a phylogenetic tree using the FastTree (v2.1.7)^51^ software with the flags --nt (input is nucleotide alignment), --gtr (generally time-reversible model) and --gamma (for branch-length rescaling and calculation of gamma20-likelihood).

### Statistics for cohort comparisons

Basal phenotypes of age and BMI were compared between cohorts using pairwise Wilcoxon rank sum test using the R-base function *pairwise.wilcox.test()* and the default method “holm” for *p*-value correction. Within sample diversity was assessed using the total number of observed genera and Shannon diversity index calculated on genus level using the vegan^52^::*diversity*() function in R. To generate Shannon genus level equivalents, the Shannon diversity was used as exponent in the natural exponent function exp(). Differences between cohorts were assessed using a pairwise Wilcoxon rank-sum test implemented in the R-base function p*airwise.wilcox.test()* and the default method “holm” for *p*-value correction. Pairwise cohort differences in between sample diversity (beta diversity) were assessed using genus-level Bray-Curtis dissimilarity and a permutational multivariate analysis of variance using distance matrices as implemented in the vegan::*adonis()* function. For each comparison, 1,000 permutations were used to assess *p*-values.

### Statistical framework for genome wide association analysis

**Rationale:** The assembly of intestinal microbial communities is a highly complex process, which potentially can be driven by environmental and lifestyle factors, host-genetics^5-10^ and disease.^1-4^ These biotic and abiotic factors mould niches for specific microorganisms, supplying them with metabolic substrates which can be directly host-derived, as with specific glycosylation patterns, or influenced by the host’s metabolism, as it is discussed for the connection between the persistence of lactose hydrolysis and the abundance of Bifidobacterium.^11^ The univariate statistical frameworks applied in this study aimed to identify genetic associations with presence/absence and abundance patterns of microbial clades. These associations could be the result of variation in host genes leading to the availability of specific energy sources or metabolic substrates (and the lack thereof, respectively). Such effects would, therefore, facilitate competitive (dis-)advantage of the specific bacteria associated to them. Alternatively, an immune response, which is specific to a given microbial feature, could be influenced by genetic variations. In this case, the abundance or the presence of the microorganism in the community would be modulated. In addition, the community as a whole can be influenced by the effect of host genetics, which can act on more than a single clade, and can also depend upon stochastic effects in the initial community assembly.^53^ As such, effects would be distributed across multiple features or clades with only small individual effect sizes. Therefore, an association analysis targeting multivariate effects was additionally implemented to identify host-genetics associated shifts on the level of the microbial community.

**Feature filtering:** All univariate microbial features, defined by either taxonomic annotation or ASV/OTU clustering, independently underwent filtering using the same criteria for inclusion in the association analysis. Within a cohort, a feature had to be present in at least 100 individuals and had to exceed the median abundance of 50 reads, thus .5%, in the individuals with non-zero counts. For the analysis of differential prevalence, the feature additionally had to be absent in at least 100 individuals. If these criteria were fulfilled in at least three of the cohorts, the feature was included in the analysis. Summary statistics for all cohorts and microbial features included in the analysis can be found in **Supplementary Table S1**. This filtering resulted in 233 univariate features for the abundance-based analysis, of which were four on phylum level, eight on class level, six on order level, 10 on family level, 29 on genus level and 65, 62 and 49 on 97%-OTU, 99%-OTU and ASV level, respectively. For the presence-absence-based analysis, 198 features were included, of these two were on class, one on order, two on family, 17 on genus and 65, 62 and 49 on 97%-OTU, 99%-OTU and ASV level, respectively. In total, 431 univariate microbial features were included in the genome-wide association analysis.

**Prevalence-based analysis:** For the analysis of genetic effects on the prevalence of bacterial features, abundance values were recoded into 0 (absence) and 1 (presence). Genetic variants were filtered to a minor allele frequency of > 5% and coded into numeric features 0 (homozygous for reference allele), 1 (heterozygous) and 2 (homozygous for alternative allele). Taxon prevalence was submitted to a logistic regression employing a generalized linear model with binomial distribution and logit-link-function using the genotype as predictor, including age, sex, body mass index (BMI), and the ten first genetic principle components (PCs) as covariates. All tests statistical tests were performed two-sided.

**Abundance-based analysis:** For calculating the effects of genetic variants on the zero-truncated abundance of bacterial features, the features were first filtered for extreme outliers, deviating more than 5 interquartile ranges (IQR) from the median abundance. Using the *glm.nb()* function from the MASS package in R, count abundances were fit in a model using previously mentioned covariates age, sex, BMI and the first ten genetic PCs as covariates. Residual variation was extracted using the *residuals()* function and submitted to a linear model estimating the effect of the genetic variants on the residual abundance. Analysis of SNP vs. feature abundance directly using generalized linear models with negative binomial distribution was tested as well; however, these models’ results showed highly inflated λ_GC_-values, thus were discarded for the genome-wide association analysis. All tests statistical tests were performed two-sided.

**Beta diversity analysis:** In addition to the single-feature based analyses, we analyzed the effects of genetic variants on the beta-diversity. For this, the genus-level abundance tables were used to calculate the pairwise Bray-Curtis dissimilarity between the individual microbial communities. Additionally, weighted, normalized UniFrac distance was calculated based on 97% identity OTU abundances using the *UniFrac()* function in phyloseq. Distance-matrices were submitted to a distance-based redundancy analysis (dbRDA) using the vegan::*capscale()* function and the same previously mentioned covariates. The residual variance of the model was extracted using the *residuals()* function, resulting in a distance matrix adjusted for these possibly confounding factors. This distance matrix was used in a procedure to estimate the effect of genetic variants based on a distance-based F-test using moment matching^54^. The calculations were implemented to run on a GPU for further speed-up, especially in the larger cohorts (see supplemental data for benchmark). As calculations for large cohorts with n > 1,000 individuals (with tables of size n×n) still could not be finished in reasonable time, we employed a stepwise calculation of results for the cohorts (estimating from single CPU usage, processing time of 7×10^6^ variants for the SHIP-Trend dataset would take 61 years; and even using one GPU instance, processing would take ~94 days). The stepwise calculation process was as follows: For the PopGen, FoCus and SHIP cohort, all variants were tested for an association. If a variant showed a nominal significant association *(p* < 0.05) in at least one of the cohorts, this variant was tested in the KORA cohort. If then a variant was nominal significant in at least two of these four cohorts, it was also tested in the SHIP-TREND cohort.

**Meta-analysis:** Genomic inflation (λ_GC_) was assessed for all cohorts and features, and all showed values below the proposed threshold of 1.05. Results from the separate cohorts were combined using a meta-analysis framework. Prevalence- and abundance-based results were submitted to an inverse-variance based strategy, calculating effects based on effect size and variance of the respective cohorts. For the beta-diversity meta-analysis, we chose a weighing based on sample size of the respective cohorts. Both approaches were adapted from the METAL software package for GWAS meta-analysis.^55^ Criteria for the reporting of a significant association were a genome-wide significant meta-analysis *p*-value < 5×10^−8^, and nominal significance in at least two cohorts for the single-feature tests and at least three cohorts for the beta diversity analysis. As the univariate microbial features can be correlated across the different taxonomic levels in the analysis, the matSpDlite algorithm was used to estimate the effective number of independent (effective) variables across all levels based on the variance of eigenvalues of the univariate abundances and presence-absence patterns.^56,57^ This yielded 141 and 127 effective variables for the abundance-based and the prevalence-based analysis, respectively. From this, we defined a study-wide significance threshold of P < 5 × 10^-8^ / 268 = 1.866 × 10^-10^. Heterogeneity statistics – Cochran’s Q and variation across studies due to heterogeneity I^2^ – for individual variants from the presence/absence and relative abundance association analyses were calculated as described in Deeks *et al*. (2008).^58^

### Analysis of influence of blood groups and secretor status

Hurdle models were used to investigate prevalence and abundance patterns in connection with ABO blood group and secretor status. Nine models were used for analysis. Models 1 – 4 analysed the effects of the individual’s counts of A alleles, B alleles, the sum of A and B alleles and the binary status O vs. non-O, respectively. Models 5 – 8 investigated the same factors, however in interaction with FUT2 secretor status, thus only taking non-zero values when assigned as “secretor”. The last model only investigated the effects of the binary secretor status. All models included the covariates age, sex, BMI and the first ten genetic principle components, in analogy to the genome-wide association analysis. Inverse-variance weighted meta-analysis was used to combine the results into a composite result per taxon and model.

### Mendelian Randomization

Mendelian Randomization (MR) analysis was performed using the TwoSampleMR package (version 0.4.25)^25^ for R. Using the MR-Base database (mrbase.org), 41 binary traits from the subcategories “Anthropometric”, “Autoimmune / Inflammatory”, “Bone”, “Cancer”, “Cardiovascular”, “Diabetes”, “Kidney”, “Pediatric disease”, and “Psychiatric / neurological” were selected for analysis of directional effect of microbial features on these outcomes. A full list of the selection criteria, used outcome traits and the used database IDs can be found in the Supplemental Material. To ensure power and suitability of the instruments used for MR, only variants with *p*-value < 10^_5^ and F-statistics^59^ > 10 were included as exposure/instrument variables in the analysis. Remaining instruments were LD clumped to include only independent signals. Using the *power_prune*() function, the best set of instrumental variables for each trait was selected using instrument strength and sample size as selection criteria (method=2). Primary Mendelian randomization analysis was performed for sets with multiple instrument variables and single instrument variables using the inverse variance weighted analysis and Wald Test, respectively. Additional sensitivity analyses using weighted median, weighted mode and Egger regression were performed for analyses with more than two instrument variables available. Per microbial trait, a suggestive threshold was defined as *p* < 0.05/41 = 1.220×10^-3^. For study wide significance, *p*-values were adjusted using Benjamin-Hochberg FDR correction, for the resulting *q*-value the threshold was set to 0.05. For beta diversity analysis, no MR was performed, as the non-parametric test used for analysis did not include a beta value for effect size needed for MR.

## Data Availability

Cohort-level summaries of microbial feature abundances are provided in the supplemental matierial. The German mGWAS Browser application is available for local query of results from Dockerhub: https://hub.docker.com/r/mruehlemann/german_mgwas_browser_app. Due to constraints given by the written consent, participant phenotypes, genotyping and 16S rRNA gene sequencing data is available upon request from the respective biobanks:
PopGen and Focus: https://portal.popgen.de/
KORA FF4: https://epi.helmholtz-muenchen.de/
SHIP and SHIP-TREND: https://www.fvcm.med.uni-greifswald.de/dd_service/data_use_intro.php

https://epi.helmholtz-muenchen.de/

https://www.fvcm.med.uni-greifswald.de/dd_service/data_use_intro.php

http://ikmb.shinyapps.io/German_mGWAS_Browser

## Acknowledgements

We want to thank Mr Tonio Hauptmann, Ms Ilona Urbach and Ms Ines Wulf of the IKMB Microbiome Lab for excellent technical assistance. We are very thankful to Dr. Kaitlin Wade for her valuable input on the Mendelian Randomization analysis. We thank Martin Schulzky for the support in figure design. This work was supported by the Deutsche Forschungsgemeinschaft (DFG) Collaborative Research Center 1182 “Origin and Function of Metaorganisms” (DFG Grant: “SFB1182”; Project A2) and the DFG Cluster of Excellence 2167 “Precision Medicine in Chronic Inflammation (PMI)” (DFG Grant: “EXC2167”). The SHIP part of the study was supported by the PePPP-project (ESF/14-BM-A55_0045/16), and the RESPONSE-project (BMBF grant number 03ZZ0921E). SHIP is part of the Research Network Community Medicine of the University Medicine Greifswald, which is supported by the German Federal State of Mecklenburg-West Pomerania.

## Author contributions

A.F., J.F.B., M.M.L., and D.H. designed the experiment. G.H., M.La., W.L., U.V., H.V., S.W., and A.P. performed genotype and phenotype data collection. F.D., F.F. and H.V. performed data quality control and curation. C.B., M.C.R., K.H., K.N. and F.U.W. performed microbiome sample preparation, data generation and curation. M.W. and M.C.R. implemented ABO blood-group inference. M.C.R., S.D., and J.K. implemented statistical models and performed the (meta-)analysis. M.C.R., C.B., B.M.H., L.B.T., and L.M.S. curated and interpreted results. M.C.R., B.M.H. and S.D. wrote the manuscript draft with advice from C.B., A.F. and J.F.B.. All authors reviewed, edited and approved the final manuscript.

## Code availability

Microbiome data pre-processing, GWAS analysis and post-processing code is available via github: https://github.com/mruehlemann/german_mgwas_code.

## Data availability

Cohort-level summaries of microbial feature abundances are provided in the supplemental matierial. The German mGWAS Browser application is available for local query of results from Dockerhub: https://hub.docker.com/r7mruehlemann/german_mgwas_browser_app. Due to constraints given by the written consent, participant phenotypes, genotyping and 16S rRNA gene sequencing data is available upon request from the respective biobanks:

- PopGen and Focus: https://portal.popgen.de/
- KORA FF4: https://epi.helmholtz-muenchen.de/
- SHIP and SHIP-TREND: https://www.fvcm.med.uni-greifswald.de/dd_service/data_use_intro.php

